# Transplacental Transfer of SARS-CoV-2 Antibodies

**DOI:** 10.1101/2020.10.07.20207480

**Authors:** Dustin D. Flannery, Sigrid Gouma, Miren B. Dhudasia, Sagori Mukhopadhyay, Madeline R. Pfeifer, Emily C. Woodford, Jourdan E. Triebwasser, Jeffrey S. Gerber, Jeffrey S. Morris, Madison E. Weirick, Christopher M. McAllister, Marcus J. Bolton, Claudia P. Arevalo, Elizabeth M. Anderson, Eileen C. Goodwin, Scott E. Hensley, Karen M. Puopolo

**Author notes:** These authors contributed equally to this work (Dustin D. Flannery, Sigrid Gouma).

## Abstract

We measured SARS-CoV-2 antibody levels in serum samples from 1,471 mother/newborn dyads and found efficient transplacental transfer of SARS-CoV-2 IgG antibodies in 72 of 83 seropositive pregnant women. Transfer ratios >1.0 were observed among women with an asymptomatic SARS-CoV-2 infection as well as those with mild, moderate and severe COVID-19. Our findings demonstrate the potential for maternally-derived antibodies to provide neonatal protection from SARS-CoV-2 infection.

---

Newborn protection from infection is primarily dependent upon neonatal innate immune responses and maternally-derived, transplacentally-acquired antibodies. The extent to which maternal antibodies produced in response to severe acute respiratory syndrome coronavirus 2 (SARS-CoV-2) infection during pregnancy cross the placenta is important for understanding potential neonatal protection from coronavirus disease 2019 (COVID-19), and for developing appropriate maternal vaccination strategies when effective vaccines become available. To date, studies of transplacental transfer of maternal SARS-CoV-2-specific antibodies to newborns are limited to case reports and small case series of women with symptomatic infection.^1–3^

To assess the relationship between maternal and neonatal SARS-CoV-2-specific antibody levels, we utilized sera and paired cord blood collected from women presenting for delivery from April 9, 2020 to August 8, 2020 at Pennsylvania Hospital in Philadelphia, Pennsylvania. We collected samples that were otherwise to be discarded and measured maternal and neonatal SARS-CoV-2 immunoglobulin G (IgG) and immunoglobulin M (IgM) antibodies to the spike receptor binding domain (RBD) antigen using an enzyme-linked immunosorbent assay (ELISA) with ∼1% false-positive rate.^4^ A cutoff at 0.48 arbitrary units was used to define IgG and IgM seropositivity, as previously described.^4^

There were 1,714 women who delivered during the study period, including 31 women with twin deliveries. Matched maternal-cord blood sera were available for 1,471 mother/newborn dyads (**Supplemental Figure 1**). Of the 1,471 matched dyads, 83 women (5.6%) were SARS-CoV-2 seropositive and 1,388 women (94.4%) were seronegative. Among infants delivered to seropositive women, 72 infants (87%) were seropositive and 11 (13%) were seronegative. There were no seropositive infants born to the 1,388 seronegative women. During the study period, pregnant women were routinely screened for SARS-CoV-2 by nasopharyngeal PCR (NP-PCR) when admitted to the hospital for delivery, and/or tested during pregnancy due to SARS-CoV-2 exposures or COVID-19 symptoms. All seropositive women were NP-PCR tested (with the exception of one woman who declined screening on admission for delivery); 44/82 (54%) seropositive women were PCR-positive at some point during pregnancy (**Table 1**). Most women (50/83) were asymptomatic for COVID-19. Newborns (20/83) were tested for SARS-CoV-2 by NP-PCR between 24-48 hours after birth if the mother was NP-PCR positive and met clinical criteria for being contagious at the time of delivery; none were positive.

**Table 1.**
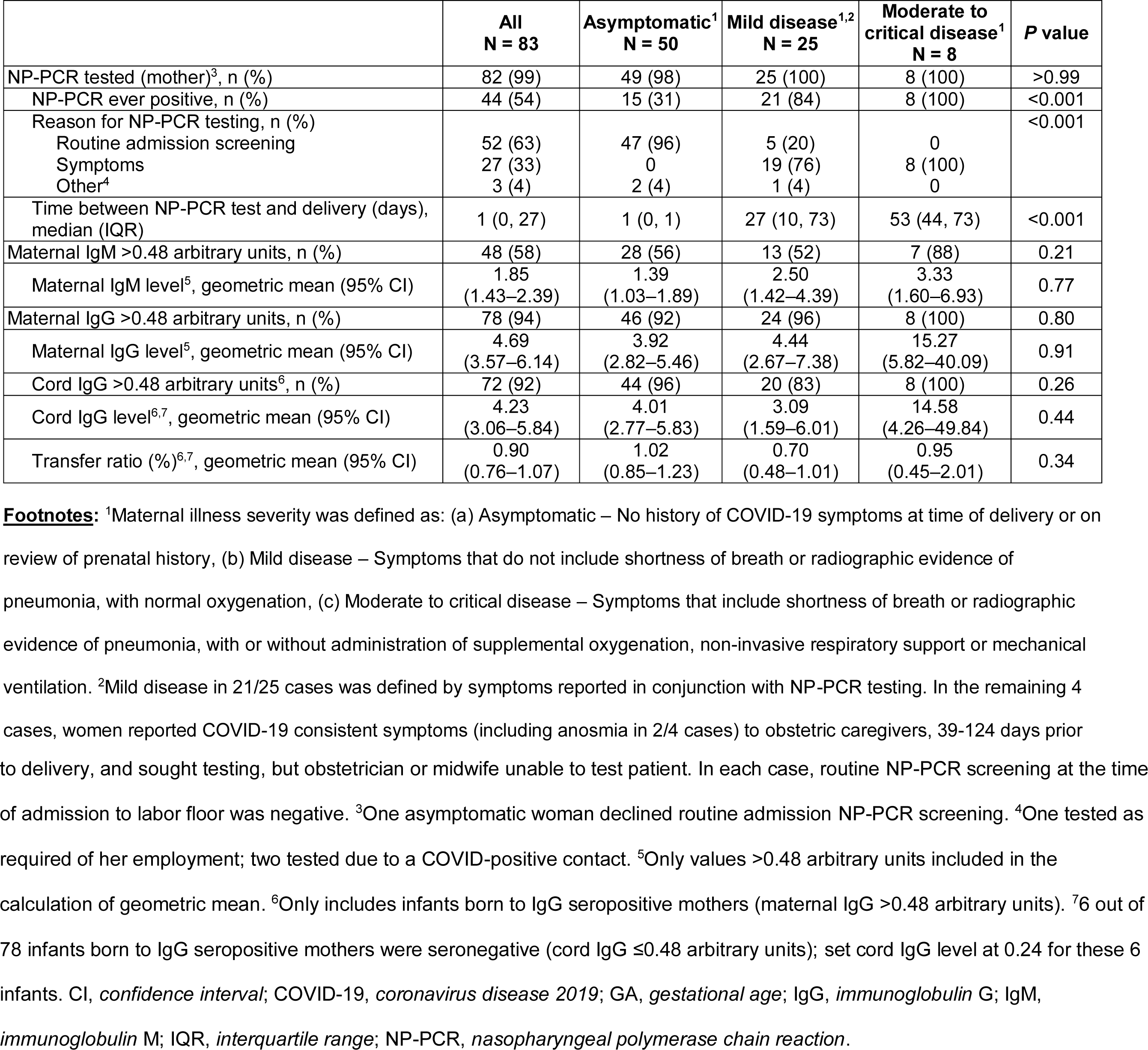
Maternal illness severity and results of NP-PCR testing and antibody levels.

|  | All<br>N = 83 | Asymptomatic<sup>1</sup><br>N = 50 | Mild disease<sup>1,2</sup><br>N = 25 | Moderate to<br>critical disease<sup>1</sup><br>N = 8 | P value |
| --- | --- | --- | --- | --- | --- |
| NP-PCR tested (mother)<sup>3</sup>, n (%) | 82 (99) | 49 (98) | 25 (100) | 8 (100) | >0.99 |
| NP-PCR ever positive, n (%) | 44 (54) | 15 (31) | 21 (84) | 8 (100) | <0.001 |
| Reason for NP-PCR testing, n (%) |  |  |  |  | <0.001 |
| Routine admission screening | 52 (63) | 47 (96) | 5 (20) | 0 |  |
| Symptoms | 27 (33) | 0 | 19 (76) | 8 (100) |  |
| Other<sup>4</sup> | 3 (4) | 2 (4) | 1 (4) | 0 |  |
| Time between NP-PCR test and delivery (days),<br>median (IQR) | 1 (0, 27) | 1 (0, 1) | 27 (10, 73) | 53 (44, 73) | <0.001 |
| Maternal IgM >0.48 arbitrary units, n (%) | 48 (58) | 28 (56) | 13 (52) | 7 (88) | 0.21 |
| Maternal IgM level<sup>5</sup>, geometric mean (95% CI) | 1.85<br>(1.43–2.39) | 1.39<br>(1.03–1.89) | 2.50<br>(1.42–4.39) | 3.33<br>(1.60–6.93) | 0.77 |
| Maternal IgG >0.48 arbitrary units, n (%) | 78 (94) | 46 (92) | 24 (96) | 8 (100) | 0.80 |
| Maternal IgG level<sup>5</sup>, geometric mean (95% CI) | 4.69<br>(3.57–6.14) | 3.92<br>(2.82–5.46) | 4.44<br>(2.67–7.38) | 15.27<br>(5.82–40.09) | 0.91 |
| Cord IgG >0.48 arbitrary units<sup>6</sup>, n (%) | 72 (92) | 44 (96) | 20 (83) | 8 (100) | 0.26 |
| Cord IgG level<sup>6,7</sup>, geometric mean (95% CI) | 4.23<br>(3.06–5.84) | 4.01<br>(2.77–5.83) | 3.09<br>(1.59–6.01) | 14.58<br>(4.26–49.84) | 0.44 |
| Transfer ratio (%)<sup>6,7</sup>, geometric mean (95% CI) | 0.90<br>(0.76–1.07) | 1.02<br>(0.85–1.23) | 0.70<br>(0.48–1.01) | 0.95<br>(0.45–2.01) | 0.34 |

**Footnotes:** <sup>1</sup>Maternal illness severity was defined as: (a) Asymptomatic – No history of COVID-19 symptoms at time of delivery or on review of prenatal history, (b) Mild disease – Symptoms that do not include shortness of breath or radiographic evidence of pneumonia, with normal oxygenation, (c) Moderate to critical disease – Symptoms that include shortness of breath or radiographic evidence of pneumonia, with or without administration of supplemental oxygenation, non-invasive respiratory support or mechanical ventilation. <sup>2</sup>Mild disease in 21/25 cases was defined by symptoms reported in conjunction with NP-PCR testing. In the remaining 4 cases, women reported COVID-19 consistent symptoms (including anosmia in 2/4 cases) to obstetric caregivers, 39-124 days prior
to delivery, and sought testing, but obstetrician or midwife unable to test patient. In each case, routine NP-PCR screening at the time of admission to labor floor was negative. <sup>3</sup>One asymptomatic woman declined routine admission NP-PCR screening. <sup>4</sup>One tested as required of her employment; two tested due to a COVID-positive contact. <sup>5</sup>Only values >0.48 arbitrary units included in the calculation of geometric mean. <sup>6</sup>Only includes infants born to IgG seropositive mothers (maternal IgG >0.48 arbitrary units). <sup>7</sup>6 out of 78 infants born to IgG seropositive mothers were seronegative (cord IgG ≤0.48 arbitrary units); set cord IgG level at 0.24 for these 6 infants. CI, *confidence interval*; COVID-19, *coronavirus disease 2019*; GA, *gestational age*; IgG, *immunoglobulin G*; IgM, *immunoglobulin M*; IQR, *interquartile range*; NP-PCR, *nasopharyngeal polymerase chain reaction*.

There was a positive correlation between SARS-CoV-2 IgG levels in cord and maternal sera (Pearson’s r^2^=0.7852, *P*<0.001; **Figure 1A**). SARS-CoV-2 IgM antibodies were not detectable in any of the 72 seropositive infants (**Figure 1B**). We compared the quantitative levels of maternal IgG among the 11 seronegative infants versus seropositive infants. In 5 cases, the mother was seropositive only by IgM (without IgG above cutoff level) and the cord sera were seronegative. In the remaining 6 seronegative infants, geometric mean IgG levels were significantly lower in the mothers of the seronegative infants compared to the 72 women with seropositive infants (1.27 vs 5.22 arbitrary units, *P*=0.005) (**Supplemental Table 1**). We assessed the relationship between severity of maternal infection, maternal IgG level and cord IgG level (**Table 1**). Women with moderate or critical illness trended to higher IgG and IgM levels, and infants born to these women trended to higher cord IgG levels, but these differences in antibody levels were not statistically significant.

**Figure 1:**
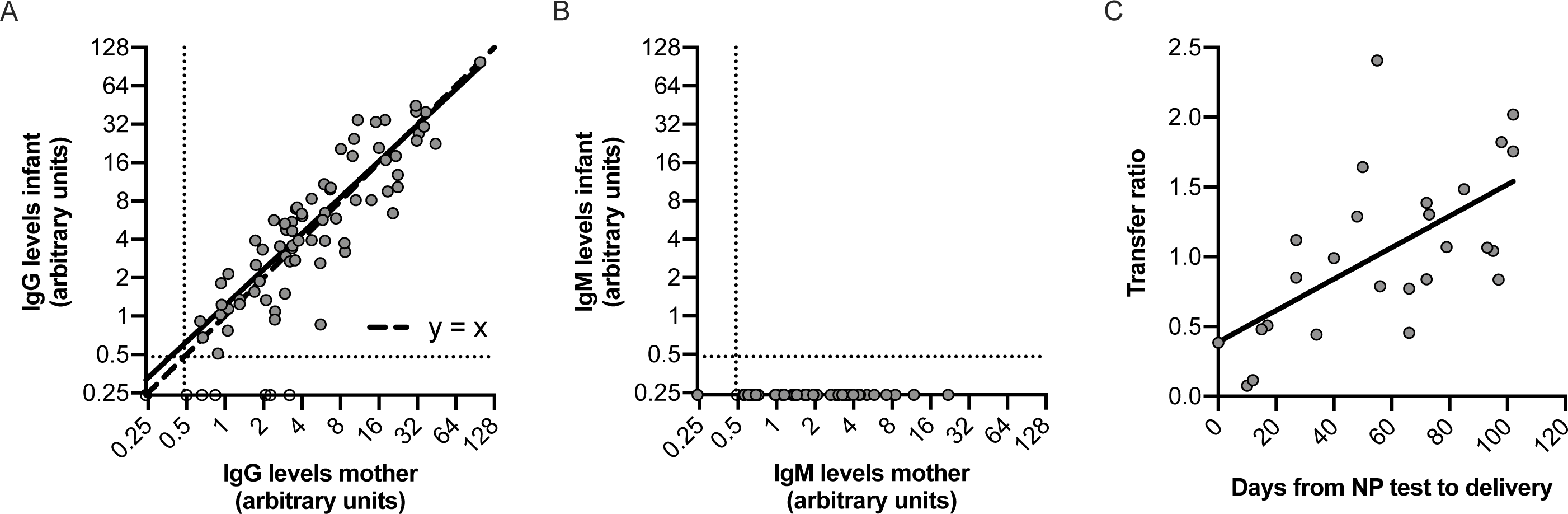
Correlation between maternal and neonatal cord sera SARS-CoV-2-specific antibody levels. (A-B) Correlation between IgG levels (A) and IgM levels (B) in sera from seropositive women and matched cord blood from seropositive (n=72, grey data points) and seronegative (n=11, open circle data points) infants. IgG levels in cord blood positively correlate with maternal IgG levels (r^2^=0.7852, *P*<0.001 using Pearson’s correlation with log_2_ transformed IgG levels).None of the infants born to seropositive mothers had detectable IgM levels. Dashed lines indicate 0.48 arbitrary units, which was the cutoff used to distinguish positive versus negative samples. Samples that were below this cutoff were assigned an antibody level of 0.24 arbitrary units. (C) Effect of days from NP test to delivery on transplacental antibody transfer. Transfer ratio of IgG antibodies from mother to infant (n=26 matched mother-infant dyads) is positively correlated with days from NP test to delivery (r^2^=0.3845, *P*<0.001 using Pearson’s correlation).

We determined the transplacental antibody transfer ratio, calculated as infant IgG level divided by maternal IgG level. Transfer ratio was not different among cord sera from infants born to mothers according to level of illness (**Table 1**). In contrast to asymptomatic women whose onset of exposure or infection cannot be reliably determined, the timing of symptom-prompted viral testing can be used as a surrogate for onset of infection. Therefore, we assessed the relationship between transfer ratio and onset of maternal infection among a subset of 26 women with mild, moderate or critical illness, who had a positive NP-PCR test prior to delivery and who delivered at term gestation. We observed a positive correlation between transfer ratio and increasing time between NP-PCR testing and delivery (Pearson’s r^2^=0.3845, *P*<0.001; **Figure 1C**). We further explored the impact of maternal, fetal and newborn characteristics on categorical cord serostatus among all seropositive mothers (n=83) and categorical transfer ratio among IgG seropositive mothers (n=78). We found no association with maternal demographic and pregnancy health characteristics and cord seropositive status (**Supplemental Table 1**). Transfer ratio was not different comparing preterm (<37 weeks’ gestation) versus term (≥37 weeks’ gestation) deliveries, with the earliest delivery at 31 weeks’ gestation (**Supplemental Table 2**).

Determination of correlates of maternal and neonatal immunity is a high priority area for SARS-CoV-2 research in the pregnancy and newborn health domains.^5^ Although placental and neonatal SARS-CoV-2 transmission have been reported^1–3,6–8^, current evidence suggests that this event is not common.^9,10^ Two case reports from China describing detectable neonatal IgM soon after birth could represent true vertical transmission of disease, or could represent false-positive IgM testing.^2,3^ We did not detect IgM antibodies in any cord blood sera samples even in cases of critical maternal illness or preterm delivery, supporting that maternal-fetal SARS-CoV-2 transmission is rare. Of greater concern is the potential for newborns to be infected postnatally from contagious mothers or other household contacts. We found efficient transplacental transfer of IgG antibodies from SARS-CoV-2 seropositive pregnant women, with transfer ratios that could exceed 2 in this study where ∼90% of women delivered at term gestation. We found that higher maternal antibody levels were associated with higher cord antibody levels. Furthermore, a longer duration between onset of maternal infection and time of delivery led to higher maternal antibody levels and higher transfer ratios. These findings are aligned with prior studies of transplacental antibody transfer, although other factors – such as the antigen-elicited IgG subclass, maternal infections during pregnancy, chronic maternal immunodeficiency, placental pathology and gestational age at birth – are known to impact transfer efficiency.^11,12^

Strengths of our study include a large cohort with access to available discarded specimens, allowing a focus on all women presenting for delivery over the study period, as opposed to studies targeting women with clinical infection identified during pregnancy or at the time of delivery. Study limitations include the small numbers of samples from preterm births, and the use of discarded specimens with retrospective data collection that does not provide information on post-discharge outcomes.

It is notable that the majority of seropositive women were asymptomatic, within uncertain timing of viral exposure. Among the subset of women in our study whose onset of infection could be estimated by symptoms prompting viral NP-PCR testing, all cord sera were seropositive if the time between maternal NP-PCR testing was at least 17 days prior to delivery. Further studies are needed to determine if SARS-CoV-2 antibodies are protective from newborn infection; if so, at what level; and whether the transplacental kinetics of vaccine-elicited antibodies are similar to naturally-acquired antibodies. Our findings demonstrate the potential for maternally-derived antibodies to provide neonatal protection from SARS-CoV-2 infection, and will help inform both neonatal management guidance as well as the design of vaccine trials during pregnancy.

## Methods

### Serum sample collection and clinical data

We utilized discarded maternal serum samples and discarded neonatal cord blood sera collected during the birth hospital admission. Pregnant women at Pennsylvania Hospital have blood drawn for rapid plasma reagin at the time of delivery as routine syphilis screening and newborns routinely have cord blood stored if blood type or Coombs testing is clinically needed; residual maternal and cord blood serum from this testing was obtained when it would otherwise be discarded. Demographic and clinical data, including information on additional SARS-CoV-2 testing prior to delivery, were collected from review of electronic medical records. Race and ethnicity were self-reported. International Classification of Diseases, 10th revision (ICD-10) diagnosis codes O24, E08-E13, and Z79.4 were used to capture type 1 diabetes, type 2 diabetes, and gestational diabetes, and codes O10, O11, O13-O16, I10-I13, and I15 were used to capture hypertensive disorders, gestational hypertension, and pre-eclampsia. The accuracy of using these codes has been validated with medical chart review^4^. Only the first twin from each pair as included in all analyses.

### Enzyme-linked immunosorbent assay (ELISA)

Samples were tested in ELISA using plates coated with recombinant receptor binding domain (RBD) of the SARS-CoV-2 spike protein. This assay has been validated using samples from COVID-19 recovered donors and samples collected prior to the COVID-19 pandemic, as previously described.^4^ The RBD protein was produced in 293F cells using plasmid provided by F. Krammer (Mt. Sinai) and purified by nickel-nitrilotriacetic acid resin (Qiagen). Coated ELISA plates (Immulon 4 HBX, Thermo Fisher Scientific) were stored overnight at 4°C. The next day, plates were washed 3 times with phosphate-buffered saline containing 0.1% Tween-20 (PBS-T) followed by blocking for 1 hour with PBS-T supplemented with 3% non-fat milk powder. Heat-inactivated serum samples were diluted in PBS-T supplemented with 1% non-fat milk powder and 50 µL diluted serum sample was added to each well after washing the plates 3 times with PBS-T. After 2 hours incubation at room temperature using a plate mixer, plates were washed again 3 times with PBS-T and 50 µL of 1:5000 diluted horseradish peroxidase labeled goat anti-human IgG (Jackson ImmunoResearch Laboratories) or 1:1000 diluted goat anti-human IgM-horseradish peroxidase (SouthernBiotech) secondary antibodies were added. Plates were incubated for 1 hour at room temperature using a plate mixer, followed by another triple wash with PBS-T. Next, 50 µL SureBlue 3,3’,5,5’-tetramethylbenzidine substrate (KPL) was added to each well and plates were incubated for 5 minutes before 25 µL of 250 mM hydrochloric acid was added to each well to stop the reaction. Plates were read with the SpectraMax 190 microplate reader (Molecular Devices) at an optical density (OD) of 450 nm. All samples were also tested on plates coated with PBS only to obtain background OD values, which were subtracted from the OD values from plates coated with RBD. Monoclonal antibody CR3022 was included on each plate to adjust for plate-to-plate variation and antibody concentrations were expressed in arbitrary units relative to CR3022. Plasmids to express CR3022 were provided by I. Wilson (Scripps). All samples were first tested in duplicate at a 1:50 dilution. Samples with an IgG and/or IgM concentration above the lower limit of detection (0.20 arbitrary units) were repeated in at least a 7-point dilution series to obtain quantitative results. Samples with IgG and/or IgM levels >0.48 arbitrary units were considered seropositive. Samples with IgG and/or IgM levels below this cutoff were assigned a value of 0.24 arbitrary units for all analyses. We previously reported IgG levels for a maternal seroprevalence study which included 25 of the 83 seropositive women in the current report.^4^

### Ethics approval

The Institutional Review Board of the University of Pennsylvania approved this study with waiver of informed consent.

### Statistical methods

Antibody levels were log_2_-transformed for all statistical analyses. Geometric mean antibody levels with 95% confidence intervals were reported if not stated otherwise. The correlation between log_2_-transformed maternal and neonatal IgG levels as well as the correlation between transfer ratio and days between NP test and delivery were reported using Pearson’s correlation coefficient. Standard descriptive analyses using Fisher’s exact test, unpaired *t* test, analysis of variance (ANOVA), Mann-Whitney *U* test, and Kruskal-Wallis test as appropriate, compared the demographics, clinical characteristics, timing and reason for maternal NP-PCR testing, and maternal and infant antibody levels and transfer ratios between the comparison groups. Statistical significance was set at *P*<0.05. Statistical analyses were performed using Stata version 16 (StataCorp, College Station, TX) and Prism version 8 (GraphPad Software).

### Data availability

De-identified individual-level data are available upon reasonable request to SEH and KMP at.

## Data Availability

De-identified individual-level data are available upon reasonable request to SEH and KMP at.

## Abbreviations

COVID-19: coronavirus disease 2019
ELISA: enzyme-linked immunosorbent assay
ICD-10: International Classification of Diseases, 10^th^ revision
IgG: Immunoglobulin G
IgM: Immunoglobulin M
IUGR: intrauterine growth restriction
NP: nasopharyngeal
OD: optical density
PBS-T: phosphate-buffered saline supplemented with Tween-20
PCR: polymerase chain reaction
RBD: receptor binding domain
SARS-CoV-2: severe acute respiratory syndrome coronavirus 2

## Acknowledgments

We thank Jeffrey Lurie and Joel Embiid, Josh Harris and David Blitzer for philanthropic support. We thank Florian Krammer (Mt. Sinai) for providing the SARS-CoV-2 spike RBD expression plasmids and Ian Wilson (Scripps) for providing plasmids to express monoclonal CR3022.

Funding for this study was provided in part by a Children’s Hospital of Philadelphia Foerderer Grant for Excellence to KMP.

## Author Contributions

DDF conceptualized and designed the study, collected data, drafted the initial manuscript, and revised the manuscript.

SG conceptualized and designed the study, led the serological experiments, collected data, drafted the initial manuscript, and revised the manuscript.

MBD designed the data collection instruments, collected data, carried out the analyses, and revised the manuscript.

SM conceptualized and designed the study, designed the data collection instruments, carried out the analyses, and revised the manuscript.

MRP collected specimens, collected data, and revised the manuscript.

ECW collected specimens, collected data, and revised the manuscript.

JSG conceptualized and designed the study and revised the manuscript.

JET conceptualized and designed the study and revised the manuscript.

JSM conceptualized and designed the study, carried out the analyses, and revised the manuscript.

MEW completed serological assays and revised the manuscript.

CMM completed serological assays and revised the manuscript. MJB completed serological assays and revised the manuscript.

CPA completed serological assays and revised the manuscript.

EMA completed serological assays and revised the manuscript.

ECG completed serological assays and revised the manuscript.

SEH conceptualized and designed the study, coordinated and supervised serological studies, and revised the manuscript.

KMP conceptualized and designed the study, coordinated, supervised and contributed to data collection, and revised the manuscript.

---

**Supplemental Table 1.** Characteristics of seropositive mothers and their newborns (N = 83 paired maternal and neonatal cord sera)

| Characteristics | Infant seropositive<sup>1</sup><br>N = 72 | Infant seronegative<br>N = 11 | P value<sup>2</sup> |
| --- | --- | --- | --- |
| Maternal age (years), median (IQR) | 29 (25, 33) | 28 (22, 32) | 0.33 |
| Maternal race<sup>3</sup>, n (%) |  |  | 0.17 |
| Black/Non-Hispanic | 32 (44) | 4 (36) |  |
| White/Non-Hispanic | 10 (14) | 2 (18) |  |
| Asian | 1 (1) | 1 (9) |  |
| Hispanic | 28 (39) | 3 (27) |  |
| Other | 1 (1) | 1 (9) |  |
| Gravidity, n (%) |  |  | 0.67 |
| 1 | 13 (18) | 1 (9) |  |
| 2 | 17 (24) | 4 (36) |  |
| ≥3 | 42 (58) | 6 (55) |  |
| Pre-pregnancy BMI<sup>4</sup>, n (%) |  |  | 0.17 |
| <18.0 | 3 (4) | 0 |  |
| 18.5 to <25.0 | 18 (25) | 6 (55) |  |
| 25.0 to <30.0 | 26 (36) | 4 (36) |  |
| ≥30.0 | 25 (35) | 1 (9) |  |
| Diabetes<sup>5</sup>, n (%) | 6 (8) | 0 | >0.99 |
| Hypertension<sup>5</sup>, n (%) | 15 (21) | 2 (18) | >0.99 |
| Multiple gestation, n (%) | 0 | 2 (18) | 0.02 |
| Cesarean delivery, n (%) | 17 (24) | 2 (18) | >0.99 |
| GA at delivery (weeks), median (IQR) | 39 3/7<br>(38 0/7, 40 0/7) | 39 1/7<br>(37 3/7, 39 5/7) | 0.39 |
| Preterm delivery at GA <37 weeks, n (%) | 7 (10) | 2 (18) | 0.34 |
| Birth weight (grams), median (IQR) | 3315<br>(2940, 3625) | 2990<br>(2500, 3200) | 0.07 |
| APGAR score at 5 minutes <5<sup>6</sup>, n (%) | 1 (1) | 0 | >0.99 |
| Time between maternal sera sampling and delivery (hours), median (IQR) | 9 (4, 20) | 14 (6, 17) | 0.92 |
| NP-PCR tested during pregnancy, n (%) | 71 (99) | 11 (100) | >0.99 |
| NP-PCR positive, n (%) | 38 (54) | 6 (55) | >0.99 |
| Time between positive NP-PCR test and delivery (days), median (IQR) | 34 (1, 72) | 6 (0, 12) | 0.04 |
| Mother IgG seropositive<sup>1</sup>, n (%) | 72 (100) | 6 (55) | <0.001 |
| Maternal IgG level<sup>7</sup>, geometric mean (95% CI) | 5.22<br>(3.97–6.87) | 1.27<br>(0.57–2.82) | 0.005 |
| Mother IgM seropositive<sup>1</sup>, n (%) | 41 (57) | 7 (64) | 0.75 |

**Footnotes:** <sup>1</sup>Seropositivity was based on either IgG or IgM level of >0.48 arbitrary units.
<sup>2</sup>Difference in maternal age, GA at delivery (weeks), birth weight (grams), time between maternal sera sampling and delivery (hours), and time between positive NP-PCR test and
delivery (days) was tested using Mann-Whitney *U* test; difference between maternal IgG level was tested using unpaired *t* test on log<sub>2</sub>-transformed maternal IgG values; differences in proportions of all other characteristics were tested using Fisher's exact test. <sup>3</sup>Race/ethnicity was abstracted from documentation at the time of admission and in clinical practice and is usually self-reported. <sup>4</sup>Pre-pregnancy BMI was abstracted from documentation in the medical record or from patient's self-reported entry in birth registration. <sup>5</sup>Diagnoses were based on delivery admission International Classification of Diseases, 10<sup>th</sup> Revision diagnosis codes for diabetes (O24, E08-E13, and Z79.4), and hypertension (O10, O11, O13-O16, I10-I13, and I15). <sup>6</sup>APGAR score at 5 minutes was missing for 2 seropositive infants. <sup>7</sup>Only values >0.48 arbitrary units included in the calculation of geometric mean. BMI, *body mass index*; CI, *confidence interval*; GA, *gestational age*; IgG, *immunoglobulin G*; IgM, *immunoglobulin M*; IQR, *interquartile range*; NP-PCR, *nasopharyngeal polymerase chain reaction*.

**Supplementary Table 2.** Characteristics across transfer ratio categories (N = 78)

| Characteristics | No transfer<br>N = 6 | Transfer Ratio<sup>1</sup> |  |  |  |  |
| --- | --- | --- | --- | --- | --- | --- |
|  |  | <0.50<br>N = 8 | 0.50 to <1.00<br>N = 24 | 1.00 to <1.50<br>N = 19 | 1.50 to <2.00<br>N = 14 | ≥2.00<br>N = 7 |
| Preterm delivery at GA <37 weeks, n (%) | 1 (17) | 2 (25) | 2 (8) | 3 (16) | 0 | 0 |
| Maternal IgG level, geometric mean (95% CI) | 1.27<br>(0.57–2.82) | 7.11<br>(3.52–14.33) | 7.74<br>(4.45–13.48) | 3.43<br>(1.83–6.44) | 4.11<br>(2.74–6.15) | 4.82<br>(1.82–12.75) |
| Maternal IgM level<sup>2</sup>, geometric mean (95% CI) | 0.44<br>(0.16–1.18) | 1.73<br>(1.13–2.66) | 1.44<br>(0.78–2.64) | 0.57<br>(0.33–0.99) | 0.38<br>(0.25–0.57) | 0.53<br>(0.19–1.51) |
| NP-PCR tested during pregnancy, n (%) | 6 (100) | 8 (100) | 23 (96) | 19 (100) | 14 (100) | 7 (100) |
| NP-PCR positive<sup>2</sup>, n (%) | 5 (83) | 8 (100) | 11 (48) | 11 (58) | 5 (36) | 3 (43) |
| Time between positive NP-PCR test and delivery (days), median (IQR) | 10 (0, 12) | 4 (0, 26) | 27 (2, 66) | 72 (27, 85) | 50 (0, 98) | 55 (1, 102) |

**Footnotes:** <sup>1</sup>Transfer ratio calculated as (cord IgG level)/(maternal IgG level). <sup>2</sup>After excluding the 6 infants with no transfer, there was a significant difference between the transfer ratio categories for geometric mean maternal IgM level ( $P=0.01$ ) and number of NP-PCR positive women ( $P=0.04$ ); all other characteristics were not significantly different ( $P\geq 0.05$ ) between the transfer ratio categories. CI, *confidence interval*; GA, *gestational age*; IgG, *immunoglobulin G*; IgM, *immunoglobulin M*; NP-PCR, *nasopharyngeal polymerase chain reaction*; IQR, *interquartile range*.

**Supplemental Figure 1:**
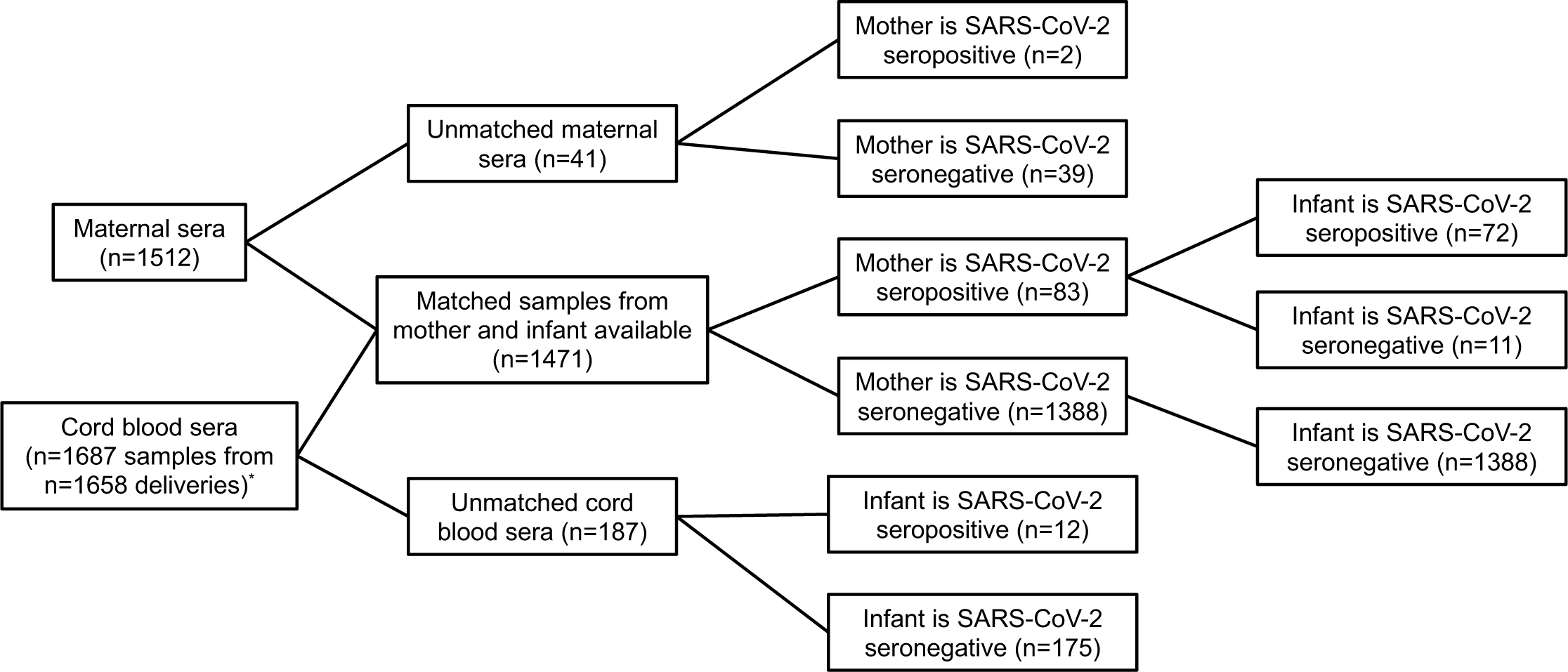
Study Flow Diagram. SARS-CoV-2 (severe acute respiratory syndrome coronavirus 2) *Includes 29 sets of twins; only one twin is included in all analyses.

